# An Individually Tailored, Home-Based Supervised Exercise Programme for People with Early Dementia: An RCT-informed Health Economic Evaluation

**DOI:** 10.1101/2024.12.10.24318781

**Authors:** Victory ‘Segun Ezeofor, Ned Hartfiel, Kodchawan Doungsong, Sarah Goldberg, Veronika van der Wardt, Louise Howe, John Gladman, Rowan H Harwood, Rhiannon T. Edwards

## Abstract

**Background:** The effectiveness of exercise interventions to improve activities of daily living function in people with dementia is inconclusive. This study aimed to assess the long-term cost-effectiveness of the PrAISED intervention from a National Health Service (NHS) perspective.

**Method:** This novel robust economic analysis used a Markov model to evaluate the incremental cost-effectiveness ratio (ICER) over a lifetime horizon of 15 years for a cohort of patients. Sensitivity analyses were conducted to investigate the uncertainty and robustness of high-impacting parameters and results.

**Results:** This study included 365 adults, aged 65 years and above with 183 and 182 randomised to the PrAISED and standard care groups respectively. The PrAISED intervention had mean per-patient cost of £60,465 for the PrAISED arm and £54,604 for standard care. The Praised intervention gained an incremental QALYs of 0.05 resulting in an ICER of £129,614 per QALY. The sensitivity analysis of the intervention cost varied the ICER value between £68,173 and £191,054/QALY. To achieve the recommended NICE willingness to pay threshold value of less than £30,000/QALYs would require the intervention cost to be reduced from £1,236 (current cost) to £263 to break even and be cost-effective. The sensitivity analyses revealed that there was a 40% probability of standard care dominating the PrAISED treatment.

**Conclusions:** Although the PrAISED intervention was a low-cost intervention, it did not produce a cost-effective intervention in this analysis. The flexibility of the PrAISED program to adapt to government policy during the COVID-19 pandemic was positive.

**Trial registration:** ISRCTN15320670

## Introduction

Dementia is a progressive condition in which there is deterioration of cognitive capacity that eventually compromises independent living ^1^. In 2019 it was estimated that over 50 million individuals globally had dementia, which is forecast to increase to 152 million by 2050^2^. There was geographical heterogeneity in the projected increases across countries and regions, with the smallest percentage changes in the number of projected dementia cases in high-income Asia Pacific (53%) and Western Europe (74%), the largest in North Africa and the Middle East (367%) and eastern sub-Saharan Africa (357%) ^3^. In the United Kingdom (UK) approximately 850,000 people currently live with dementia of whom 676,000 live in England ^4^.

The global economic costs of dementia were estimated at US$1313 billion in 2019 with high-income countries accounting for 74% of this cost ^5^. This cost superseded the estimated increase by 35% in 2015 of US$818 billion to US$1 trillion by 2018 ^6^. The estimated annual total national costs of dementia ranged from US$1.04 million in Vanuatu to US$195 billion in China ^7^. The total annual cost of dementia in England is estimated to be £24.2 billion in 2015, of which 42% (£10.1 billion) is attributable to unpaid care ^8^. Social care costs (£10.2 billion) are three times larger than health care costs (£3.8 billion), £6.2 billion of the total social care costs are met by users themselves and their families, with £4.0 billion (39.4%) funded by the government ^8^. The estimated total annual cost per person with dementia in Europe is on average €32,506.73, whereas for the United States, it gets €42,898.65 ^9^. The average total national expenditure on dementia estimated as a proportion of GDP in low and middle-income countries was 0.45%, the indirect costs, on average, accounted for 58% of the total cost of dementia, while direct costs contributed 42% ^7^.

In most studies, the life expectancy for individuals diagnosed with Alzheimer’s disease (AD) is about 7–10 years for patients in their 60s and early 70s, but findings for other types of dementia have been inconsistent ^10^. In a study by Mölsä et al, ^11^ it showed that the 14-year survival rate for AD was 2.4% versus an expected rate of 16.6%.

Regular physical exercise and activity are promising non-pharmacological interventions that could slow deterioration in cognitive function and improve activities of daily living (ADL) ^12,13^. However, the evidence is inconclusive with studies showing different results depending on the components of the exercise program. Trials of structured exercise programmes and community occupational therapy (DAPA, COTiD-UK, and EVIDEM-E) have not established a slowing of cognitive impairment in people with mild to moderate dementia and the related economic evaluations showed that these programmes are not cost-effective ^14–16^.

To establish whether physical activity and exercise can reduce the decline in activities of daily living for people with mild dementia, the UK National Institute for Health Research funded a 7-year programme of research to develop and evaluate a rehabilitation intervention (the Promoting Activity, Independence, and Stability in Early Dementia programme, PrAISED). There is little research on how to make these interventions work for with people with memory problems. The PrAISED intervention programme was a specially designed dementia-specific rehabilitation programme focussing on strength, balance, physical activity and performance of ADL, which was individually tailored to people with dementia. It provided up to 50 therapy sessions over 12 months. The control group received usual care plus a falls risk assessment ^17,18^.

This health economic evaluation aims to investigate the cost-effectiveness of individually tailored exercise in promoting activity, independence, and stability in early dementia (PrAISED) intervention compared to standard care (SC) beyond the trial period up to a lifetime horizon of 15 years.

## METHODS

### Study design

A model-based cost-utility analysis was conducted from a National Health Service (NHS) perspective to compare the PrAISED intervention programme and standard care (SC). The primary outcome measure was the incremental cost-effectiveness ratio (ICER) ^19^ which investigated the mean costs and Quality Adjusted Life Years (QALYs) over a lifetime horizon of 15 years. A 15-year lifetime horizon was applied to cover the maximum life expectancies observed in the literature.

### Decision analytic model

A six-stage age-dependent Markov model ^19,20^ was developed as a simplified clinical pathway reflection of the dementia disease severity progression (Figure1). The disease severity was based on the Activities of Daily Living (ADL), this was measured using the Disability Assessment for Dementia (DAD) ^21^. The health states were classified using the DAD scores: No or Minimal Impairment with scores of 37 – 40 out of 40, Mild Level with scores of 28 – 37 out of 40, Moderate Level with scores of 19 – 28 out of 40, Severe Impairment with scores of 9 – 19 out of 40, Very Severe Impairment with scores of 0 – 9 out of 40, and Dead: 0 out of 40. This classification was stratified by the experienced PrAISED clinicians. See Appendix A for more details.

**Figure 1:**
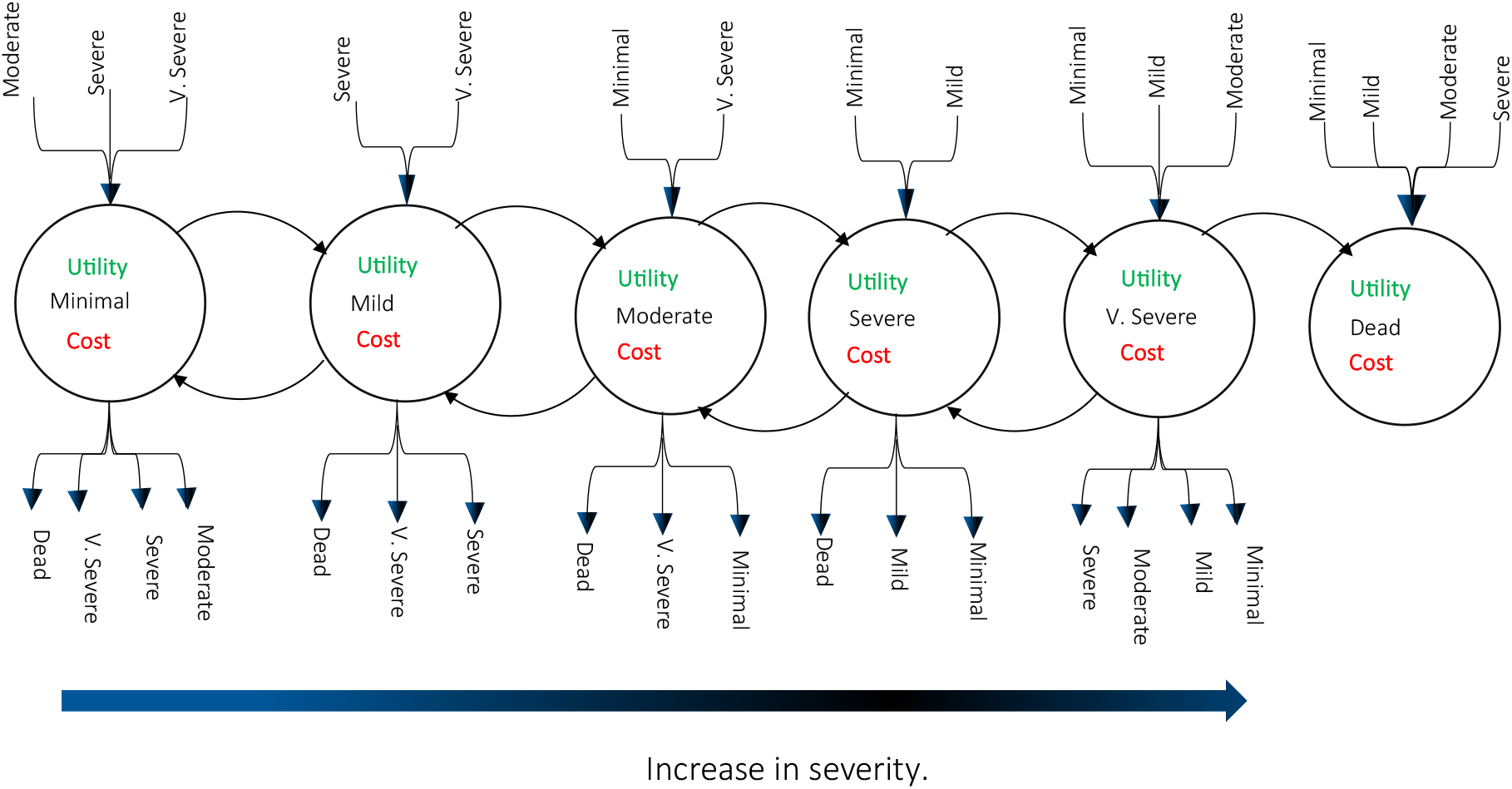
Simplified representation of the disease pathway. V. Severe = Very Severe. No or Minimal Impairment: 37 – 40 out of 40 or 92.5 – 100%, Mild Level: 28 – 37 out of 40 or 70 – 92.5%, Moderate Level: 19 – 28 out of 40 or 47.5 – 70%, Severe Impairment: 9 – 19 out of 40 or 22.5 – 47.5%, Very Severe Impairment: 0 – 9 out of 40 or 0 – 22.5%, Dead: 0 out of 40 or 0%.

Each Markov health state was attached with estimated cost, and health outcome measures (utility value); transition probabilities between the health states were also indicated to show the movement of the cohort patients from one health state to another ^22^. The cycle length was set to 12 months.

All patients are assumed to enter the system from the no or minimal health state and then transit based on the disease severity. The transition probability was obtained from the PrAISED trial data. The half-cycle correction was not applied due to the long-time horizon and the sensitivity analysis conducted, improvements in the robustness of the result due to the half-cycles were expected to be negligible.

The analysis was performed using the IBM SPSS version 25 and Microsoft Excel Office 365 software packages. This novel and robust economic model using the ADL for health state is the first and will provide evidence for future studies.

### Intervention

The PrAISED programme is a complex intervention, therefore the analysis applied was conducted in accordance with the Medical Research Council guidelines for complex interventions. It was developed as a multicomponent intervention that was dementia-specific, theoretically considered, evidence-based and feasible for and acceptable to people with mild dementia or mild cognitive impairment ^18^.

The PrAISED trial was a multi-centre, pragmatic, parallel group, randomised controlled trial, which was conducted between October 2018 and June 2022. The intervention was co-designed with stakeholders including people with dementia, carers, practitioners and policymakers ^17,18^. People aged 65 years and above with mild dementia or mild cognitive impairment living at home and a family member or carer were recruited into the trial, see Table 1. Participants in the intervention arm received an individually tailored programme comprising physical exercises to build strength and balance ^23^; functional activities to enhance ADL ^23,24^; inclusion in community life; risk enablement; and environmental assessment (accessibility and safety issues at home). The control intervention consisted of a falls prevention assessment and advice and was modelled on usual falls prevention care. For the complete details of the intervention, inclusion criteria, patient demographics and outcome measures see Bajwa et al., 2019 and Harwood et al., 2023 paper ^17,24^.

**Table 1:**
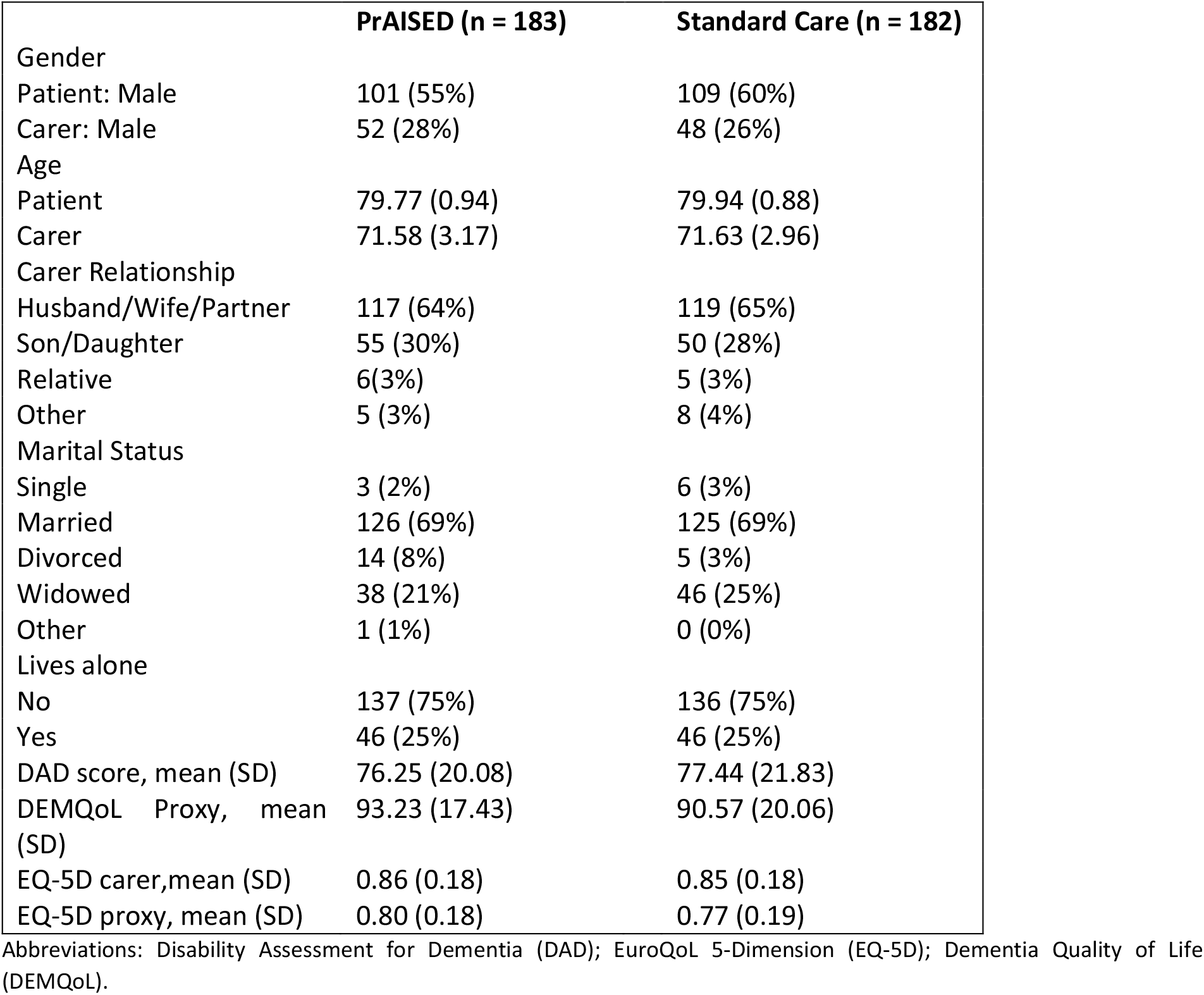
Characteristics of patient population for the PrAISED trial, n (%).

The study plan had to be adjusted with UK government’s social distancing laws during the COVID-19 pandemic. From March to September 2020, recruitment was paused; the intervention was delivered remotely via telephone or video calls, where possible, and follow up data was collected remotely via post and telephone. To understand the impact of these restrictions, the analysis included subgroups classified as:

1. During COVID-19 Pandemic: Patients who had received the PrAISED intervention via a combination of face to face and remotely during lockdown.
2. Pre-COVID-19 Pandemic: Patients who had both baseline and follow up intervention using the face-to-face interaction.
3. Total Sample: These will be the major group of this study. All patients involved in this study, irrespective of the medium they received the intervention.

The intervention delivered was the same irrespective of delivery process adopted due to the COVID-19 pandemic.

### Utilities

The unit of utility measure was the quality adjusted life-year (QALY) derived from the EQ-5D proxy. The EQ-5D-5L scores were cross-linked to the EQ-5D-3L value sets as recommended by the National Institute for Care and Excellence (NICE) ^25^. Each health state was assigned a utility value obtained from the baseline patient dataset (Table 1) which matched with the levels of dementia severity, see Appendix Table B.1 for more details.

### Transition probabilities of the Markov model

At each health state, the percentage of participants who progressed to another health state was captured as probabilities, these details are summarised in Appendix. For this health economic evaluation constant transition probabilities were applied for each cycle length.

### Mortality

Age and sex-specific death rates were obtained from the 2018-2020 UK life expectancy from the Office for National Statistics (ONS) life tables ^26^. The dementia mortality rate was obtained from the PrAISED study data.

### Data cleaning and analyses

Missing data were computed using the Markov Chain Monte Carlo (MCMC) Method of multiple imputation ^27,28^ and the data pooled via the output management system (OMS) using the IBM SPSS software.

### Treatment costs

Two components were adopted to collate the details of the cost estimate: the intervention cost, and the health resource use cost. All costs considered were analysed from a NHS perspective using the Personal Social Service Research Unit (PSSRU) Unit Costs of Health and Social Care and NHS pay ^29,30^, see Appendix C. Where costs were not identified a literature search was conducted. All costs were inflated to the 2021 values and evaluated in British Pounds Sterling (£).

Intervention costs was largely composed of staff time and were estimated with details of the resource items required with providing the PrAISED exercise intervention programme. The resource use cost was obtained using a modified client service receipt inventory (CSRI) ^31^ at 3 months up to follow-up, these costs were multiplied by 4 to estimate the annual cost per disease health state applied in the Markov model. Table 2 provides a breakdown of intervention costs used in this health economic evaluation. For more details on costs see Appendix C.

**Table 2:**
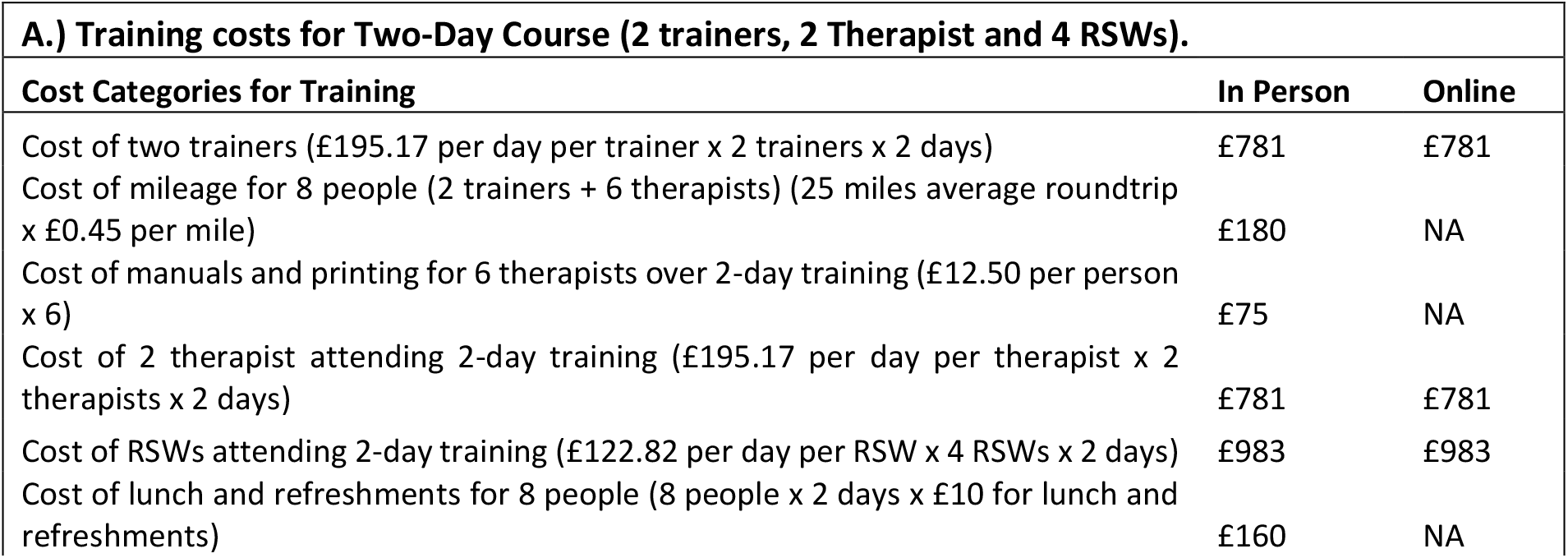
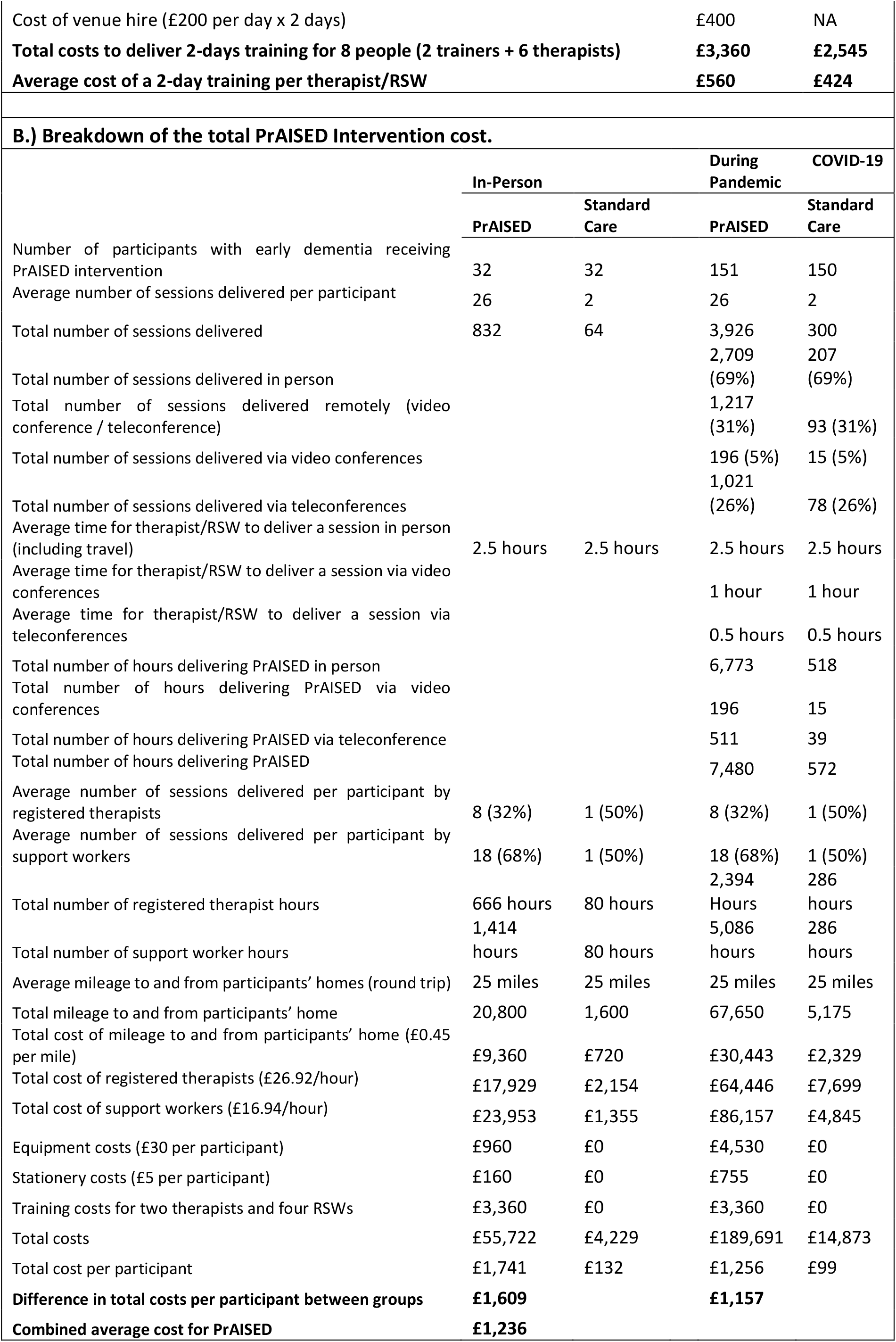
A.) Training costs for Two-Day Course for 2 trainers, 2 Therapist and 4 rehabilitation support workers (RSWs). B) Breakdown of the total PrAISED Intervention cost.

### Discounting

In order to make costs and effects occurring at different times comparable, both were discounted at 3.5% as recommended by NICE ^32^ for cost-effectiveness. In sensitivity analysis, discount rates of 0% and 1.5% were also used.

### Sensitivity analysis

To evaluate the uncertainty in the model, a deterministic sensitivity analysis [53] was used to investigate high impacting parameter values by varying them within acceptable ranges and the results summarised using the tornado diagram. A probabilistic sensitivity analysis (PSA) [53] was also conducted, the input parameters were assigned appropriate probability distributions ^20^, see Appendix Table B.1. The health state costs were assigned a gamma distribution while the QALYs and probabilities were assigned a beta distribution from which a Monte-Carlo simulation of a 1000 iteration for the ICER value was conducted ^19^. The cost-effectiveness acceptability curve (CEAC) ^20^ was generated to show the probability of the PrAISED intervention being cost-effective at various willingness-to-pay (WTP) threshold ^32^. The cost-effectiveness of the PrAISED intervention was determined with reference to the NICE cost-effectiveness threshold of £20,000 to £30,000 per QALYs gained ^32^.

## RESULTS

### Base case analysis

In the base case of the cost-utility analysis, the PrAISED intervention over the lifetime horizon (15 years) for the cohort patient accrued a higher mean cost (£60,465) than the standard care intervention (£54,604). The utility value accumulated a QALY gain of 3.45 for the PrAISED intervention, while the standard care had a 3.40 QALY gain. This led to an incremental cost of £6,223 and an incremental QALY value of 0.05 that resulted in an ICER value of £129,614 per QALY gained for the lifetime horizon analysis.

The PrAISED intervention cost of £1,236 was a low intervention cost and accounted for 2% of the mean cost, other costs were composed of the resource use costs.

Cost and utility differences based on the pre-COVID (in-person) and during COVID (remotely delivered) of the PrAISED intervention showed a similar pattern in both. The mean cost for the in-person PrAISED intervention was higher which resulted in an incremental cost of £8,880. Similarly, the mean cost for the remotely delivered PrAISED intervention was higher and had an incremental cost of £4,896. The corresponding ICER values were £107,374/QALY gained for the in-person and £171,637/QALY gained for the remotely delivered PrAISED intervention see Table 3.

**Table 3:** Baseline and lifetime cost-effectiveness analysis and probability sensitivity analysis result.

| Base case cost-effectiveness analysis for the Lifetime Horizon |  |  |  |  |  |  |  |  |
| --- | --- | --- | --- | --- | --- | --- | --- | --- |
| Intervention | Mean |  | Incremental |  | ICER (£/QALY) | Probability WTP threshold |  |  |
|  | Cost | Effect | Cost | Effect |  | £20,000/QALY | £30,000/QALY | £50,000/QALY |
| Total |  |  |  |  |  |  |  |  |
| PrAISED | £60,465 | 3.45 | £5,860 | 0.05 | £129,614 | 0.09 | 0.22 | 0.36 |
| SC | £54,604 | 3.40 |  |  |  |  |  |  |
| In-Person |  |  |  |  |  |  |  |  |
| PrAISED | £78,807 | 3.60 | £8,880 | 0.08 | £107,374 | 0.03 | 0.13 | 0.31 |
| SC | £69,927 | 3.52 |  |  |  |  |  |  |
| Remotely delivered |  |  |  |  |  |  |  |  |
| PrAISED | £52,004 | 3.19 | £4,896 | 0.03 | £171,637 | 0.12 | 0.21 | 0.33 |
| SC | £47,108 | 3.16 |  |  |  |  |  |  |

Abbreviations: Standard Care (SC).

## Sensitivity analyses

### Deterministic Sensitivity Analysis

The parameters that were most influential to the model were the cost and effect. In the one-way deterministic sensitivity analyses, the PrAISED intervention cost had a significant impact on the ICER value with an increase (decrease) of 3 standard deviations, the ICER values increased to £191,054 (decrease to £68,173) per QALY gained (Appendix Table D.1). The variation of the intervention cost by 50% did not reduce the ICER value to a value less than £30,000 per QALY gained. At a variation of the intervention cost to £263, the ICER value breaks even and falls under the NICE threshold with a value of £29,967. The variation of other parameters such as health state costs and state utility did not bring the ICER values below the £30,000/QALY gained are shown in Figure 2 panel A and Appendix Table D.1.

**Figure 2:**
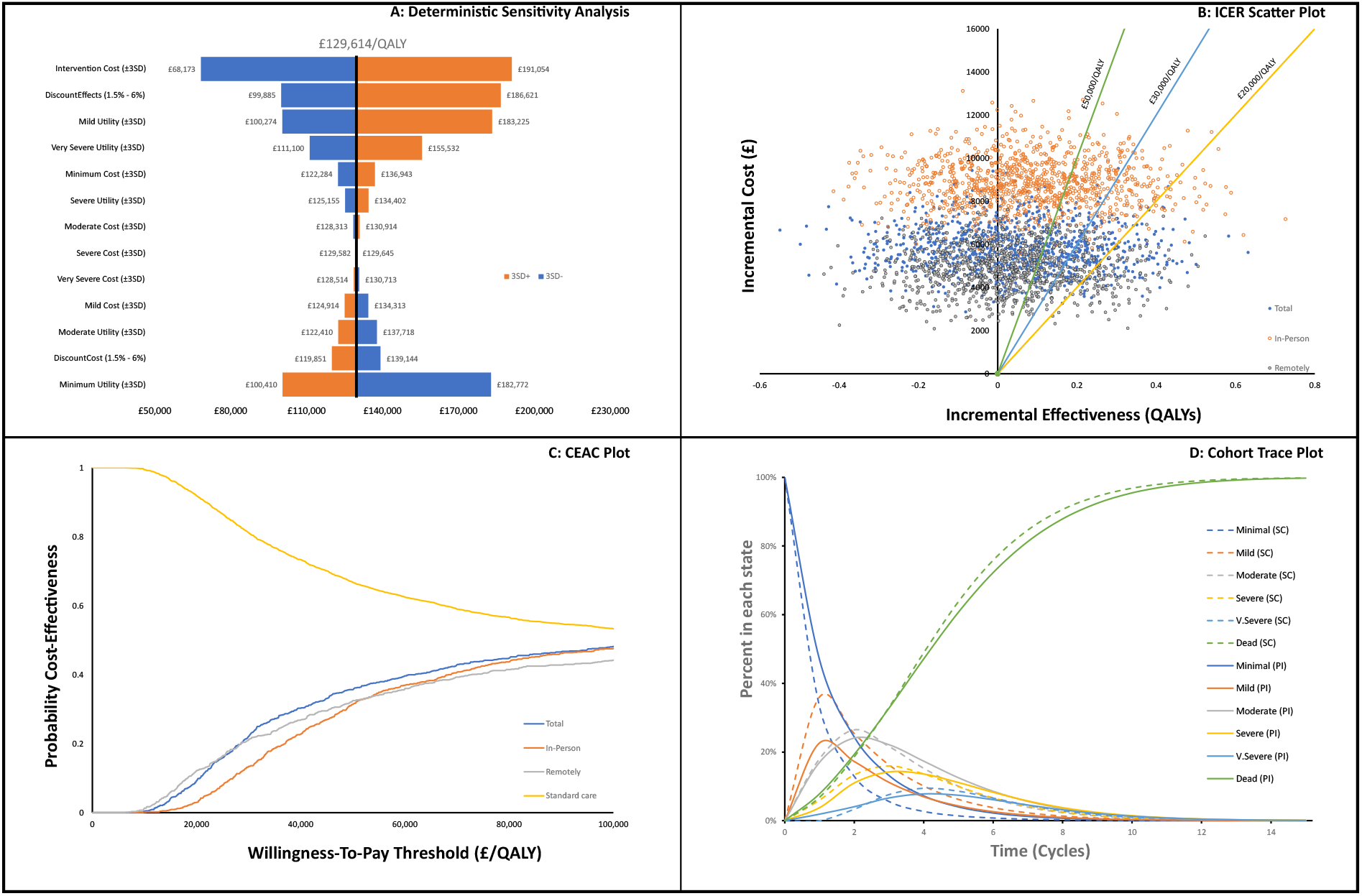
Sensitivity analysis charts: Panel A.) Deterministic sensitivity analysis, varying the intervention cost, health state cost and utility by ±3SD. SD =Standard Deviation. Panel B.) Incremental cost and Incremental effect over a lifetime comparing the PrAISED intervention to the Standard care. Panel C.) Cost-effectiveness acceptability curve for the PrAISED versus standard care intervention. Panel D.) The Markov trace shows the rate at which patients move between health states. Continuous lines. V.Severe = Very Severe; SC = Standard Care; PI = PrAISED Intervention.

### Probabilistic Sensitivity Analysis

The probability distributions of PSA net monetary benefits visually capture the distribution of a 1000 ICER simulations Figure 2 panel B, with just under 40% of the ICER values in the North-West quadrant of the plot. The scatter plot shows less than 50% of the ICER values below the £30,000/QALY. Furthermore, the CEACs shows that the PrAISED intervention at a willingness-to-pay of £20,000 per QALY had a probability of 8% being cost-effective Figure 2 panel C. At a willingness to pay threshold of £30,000 per QALY gained the PrAISED intervention had a 19% probability of being cost-effective, and it was still below the 50% probability of being cost-effective at £100,000/QALY gained.

Markov traces for the simulated cohorts transition across the model health states showed that after 7 years only 15% of the patients in the standard care arm of the trial were remaining, while 19% of patients in the PrAISED treatment arm were still alive. At the start of the 15th year (lifetime) the standard care arm had 99.76% cohort already transitioned through the model while the PrAISED treatment had 98.88% of the cohort had transitioned (Figure 2 panel D).

## DISCUSSION

The PrAISED intervention programme aimed to improve independence in ADLs and reduce the rate of functional deterioration in people with dementia or mild cognitive impairment was a low-cost intervention with a mean value of £1,236 (in-person = £1,609, hybrid (remotely delivered) = £1,157). This analysis showed a small difference in costs and QALYs between the two comparators which resulted in an incremental cost-effectiveness ratio of £129,614. The PrAISED intervention was not cost-saving for the NHS, neither did it have an ICER value below the £30,000 per QALY gained versus the standard care.

To test for uncertainty, costs and health outcomes were varied by ± 3SD from the mean values. The intervention cost being the most impactful parameter when varied was still above the NICE threshold of £30,000/QALY gained. From the PSA, the PrAISED intervention had less than 50% of the ICER iterations below the £50,000/QALY value Figure 2 panel B. The robustness of the analysis is further emphasized from the CEAC curve Figure 2 panel C which showed the same result. The trace plot (figure 2 panel D) showed that though the PrAISED intervention had lower rates of severity and mortality progression, the benefit was overcompensated with accrued cost.

### Strengths and Limitations

Modelling the effectiveness of cognitive impairment comes with its unique challenges and even more so evaluating it beyond the time period of the clinical trial. The transition probabilities were based on extrapolation of the trends shown in the data collected at baseline and follow-up periods. This model suggests that the implementation of the intervention was at all health states, and patients continued the intervention through the lifetime horizon.

One of the major strengths of this economic evaluation is that this is the first Markov model using the DAD score for the health states based on the activities of daily living (ADL). This study provides stakeholders and policy makers with evidence vital to the functional abilities of people living with dementia.

In any health economic evaluation, the health resource use cost is very important, for patients who are experiencing some degree of cognitive impairment, gathering this data is challenging ^34^. In this analysis, we had to use the information given by the carers. The resource use data was also limited to 3 months prior and up to baseline and follow-up and then multiplied by four to estimate the annual cost, this made it difficult to capture the annual cost change between the two intervention arms.

Where carers provide support to patients, the burden of the level of severity (cost and utility) might not be well captured by the patient as it will be transferred to the carer. Health economists need to look for a way to inculcate the carer strain into the utility values obtained during such trials.

The major strength of this study was that all data applied in this analysis were drawn primarily from this high quality RCT, with extensive sensitivity testing. Where uncertainty remained, these were investigated individually using the deterministic and probabilistic sensitivity analysis. The results of these analyses were robust to many changes in the parameters’ values tested.

Another strength of this study was its flexibility in being adaptable during the COVID-19 pandemic ^35^. Though being classified as a complex intervention its ability to be delivered remotely was commendable.

To further strengthen the transparency in this model, both the CHEERS checklist ^36^ and AdViSHE guideline ^37^ were strictly followed and complied with (see Appendix E).

### Comparisons with previous work

A previous economic evaluation of exercise as a therapy for behavioural and psychological symptoms of dementia within the EVIDEM-E randomised controlled trial ^16^ versus standard care, concluded that the exercise intervention did not appear to be cost-effective when considering quality-adjusted life year gains. There was a similar conclusion in the DAPA study by Khann et al. ^14^ where they investigated the cost-effectiveness of a tailored, structured, moderate-to high-intensity exercise programme versus usual care in people with mild to moderate dementia. The result presented here further deepens the understanding of the cost-effectiveness of tailored exercise interventions. The PrAISED analysis showed that the benefit obtained is minimal in comparison to the cost incurred when compared with the standard care intervention.

### Implications and Future Research and Policy

This result promotes a review of costs associated with people with mild dementia as estimated using the PSSRU cost under the NHS health care system. Our Markov modelling estimated the annual cost for patients with mild, moderate, and severe dementia at £3,988, £7348, and £12,141 – £79,753 but studies by Wittenberg et. al., ^8^ show that these costs are estimated at £21,700 – £23,000, £32,725, and £42,500 – £45,000 for the health states respectively.

Proxy measures were collected for some of the outcome measures such as the DEMQoL and EQ-5D Proxy which led to ceiling and floor effects ^38^. Finding a way in which questionnaires such as the Carer Strain Index (CSI) which measure the carers’ stress and burden can be inculcated into the QALYs of the patient will support findings on the impact of the carer.

It will be beneficial for future studies to look for exercise intervention alternatives that have lower intervention cost and provide more benefit to the patient.

The PrAISED trial used a two pronged alternative methodology to the conventional approach used in health economics, a social return on investment (SROI) ^39^ was also conducted alongside this health economic evaluation, the result challenge the deduction made here. The two divergent findings lead to the possibilities that either the conventional approach is insensitive to potential benefits whilst good at measuring costs, or that the SROIs is at risk of inflating benefits. The case for appropriateness (or not) of standard CEA in progressive, degenerative and end of life conditions is in question. This gives health service decision makers a challenge as to which methodology they find most helpful in decision making.

## CONCLUSIONS

Though this study used a novel and robust method to investigate the health economics impact of the PrAISED intervention, the analysis shows that the intervention did not offer better value for money in comparison with the standard care for patients with mild dementia or mild cognitive impairment. There was uncertainty around the mode of delivery of the PrAISED intervention due to the adaptation of the protocol during the COVID-19 pandemic, this may have influenced the study results, but there is insufficient data to demonstrate this.

## Data Availability

The datasets used and/or analysed during the current study are available from the corresponding author on request or please contact the Principal Investigator Rowan H Harwood for datasets used in the PrAISED study.

## ABBREVIATION

ADL: Activities of Daily Living
CEAC: Cost-Effectiveness Analysis Curve
CSI: Carer Strain Index
DAD: Disability Assessment for Dementia
DEMQoL: Dementia Quality of Life
EQ-5D: Euroqol-five Dimension
ICER: Incremental Cost-Effectiveness Ratio
HEAP: Health Economics Analysis Plan
NHS: National Health Service
NICE: National Institute for Care and Excellence
PSA: Probabilistic Sensitivity Analysis
QALY: Quality Adjusted Life Year
SD: Standard Deviation
WTP: Willingness-To-Pay

## Acknowledgments

The authors acknowledge the collaborations between NHS sites with respect to the intellectual property related to the PrAISED trial. We also thank the patients who participated and the public and patient involvement representatives. We want to acknowledge Dr Carys Jones for her initial work on the PrAISED HEAP. Our thanks also to Dr Catherine Lawrence, reader support for Professor Rhiannon Tudor Edwards.

## Ethical approval

Ethical approval was provided by the Yorkshire and The Humber – Bradford Leeds National Research Ethics Service (NRES) Committee Reference 18/YH/0059, with IRAS project identification 236099 and research governance departments in each organisation. All participants gave written informed consent to take part in the research.

## Author contributions

RTE reviewed the Health Economic Analysis Plan (HEAP). VE and RTE drafted the initial manuscript and VE undertook the health economics analysis and drafted the health economics manuscript; NH and KD cost the PrAISED intervention programme; LH and RH led the team in health state classification using the DAD scores; RH, JG, SG, and VvdW conceptualized the trial, designed and wrote the study protocol; All authors were involved in the drafting and reviewing of the manuscript for the final approval.

## Consent for publication

This paper presents independent research funded by the National Institute for Health Research (NIHR). The views expressed are those of the author(s) and not necessarily those of the NHS, the NIHR, or the Department of Health and Social Care.

## Consent to participate

All patients who took part in this trial consented to the study and the appropriate institutional forms have been archived.

## Code availability

The code for the health economics PrAISED analysis used in this study is available from the corresponding author upon request.

## REFERENCE

1. Iadecola C, Duering M, Hachinski V, et al. Vascular cognitive impairment and dementia: JACC scientific expert panel. J Am Coll Cardiol. 2019;73(25):3326–3344.

2. International AD. World Alzheimer Report 2019: Attitudes to dementia. Published online September 20, 2019. Accessed February 3, 2023. https://www.alzint.org/resource/world-alzheimer-report-2019/

3. Nichols E, Steinmetz JD, Vollset SE, et al. Estimation of the global prevalence of dementia in 2019 and forecasted prevalence in 2050: an analysis for the Global Burden of Disease Study 2019. Lancet Public Health. 2022;7(2):e105–e125.

4. NHS England » ementia. Accessed February 3, 2023. https://www.england.nhs.ukmental-health/dementia/

5. Wimo A, Seeher K, Cataldi R, et al. The worldwide costs of dementia in 2019. Alzheimers Dement. Published online 2023.

6. Wimo A, Guerchet M, Ali GC, et al. The worldwide costs of dementia 2015 and comparisons with 2010. Alzheimers Dement. 2017;13(1):1–7.

7. Mattap SM, Mohan D, McGrattan AM, et al. The economic burden of dementia in low-and middle-income countries (LMICs): a systematic review. BMJ Glob Health. 2022;7(4):e007409.

8. Wittenberg R, Knapp M, Hu B, et al. The costs of dementia in England. Int J Geriatr Psychiatry. 2019;34(7):1095–1103.

9. Cantarero-Prieto D, Leon PL, Blazquez-Fernandez C, Juan PS, Cobo CS. The economic cost of dementia: a systematic review. Dementia. 2020;19(8):2637–2657.

10. iang CS, i J, ang FC, et al. Mortality rates in Alzheimer’s disease and non-Alzheimer’s dementias: a systematic review and meta-analysis. Lancet Healthy Longev. 2021;2(8):e479–e488. doi:10.1016/S2666-7568(21)00140-9

11. Mölsä PK, Marttila RJ, Rinne UK. Long-term survival and predictors of mortality in Alzheimer’s disease and multi-infarct dementia. Acta Neurol Scand. 1995;91(3):159–164. doi:10.1111/j.1600-0404.1995.tb00426.x

12. Forbes D, Thiessen EJ, Blake CM, Forbes SS, Forbes S. Exercise programs for people with dementia. Sao Paulo Med J. 2014;132:195–196.

13. Bossers WJ, Van der Woude LH, Boersma F, Hortobágyi T, Scherder EJ, Van Heuvelen MJ. Comparison of effect of two exercise programs on activities of daily living in individuals with dementia: a 9-week randomized, controlled trial. J Am Geriatr Soc. 2016;64(6):1258–1266.

14. Khan I, Petrou S, Khan K, et al. Does structured exercise improve cognitive impairment in people with mild to moderate dementia? a cost-effectiveness analysis from a confirmatory randomised controlled trial: the dementia and physical activity (DAPA) trial. PharmacoEconomics-Open. 2019;3:215–227.

15. Pizzo E, Wenborn J, Burgess J, et al. Cost-utility analysis of community occupational therapy in dementia (COTiD-UK) versus usual care: Results from VALID, a multi-site randomised controlled trial in the UK. Plos One. 2022;17(2):e0262828.

16. d’Amico F, ehill A, Knapp M, et al. Cost-effectiveness of exercise as a therapy for behavioural and psychological symptoms of dementia within the EVIDEM-E randomised controlled trial. Int J Geriatr Psychiatry. 2016;31(6):656–665.

17. Bajwa RK, Goldberg SE, Van der Wardt V, et al. A randomised controlled trial of an exercise intervention promoting activity, independence and stability in older adults with mild cognitive impairment and early dementia (PrAISED)-A Protocol. Trials. 2019;20(1):1–11.

18. Booth V, Harwood RH, Hood-Moore V, et al. Promoting activity, independence and stability in early dementia and mild cognitive impairment (PrAISED): development of an intervention for people with mild cognitive impairment and dementia. Clin Rehabil. 2018;32(7):855–864.

19. Briggs A, Sculpher M. An introduction to Markov modelling for economic evaluation. Pharmacoeconomics. 1998;13(4):397–409.

20. Edlin R, McCabe C, Hulme C, Hall P, Wright J. Cost effectiveness modelling for health technology assessment. Published online 2019.

21. Gélinas I, Gauthier L, McIntyre M, Gauthier S. Development of a functional measure for persons with Alzheimer’s disease: the disability assessment for dementia. Am J Occup Ther. 1999;53(5):471–481.

22. Petrou S, Gray A. Economic evaluation using decision analytical modelling: design, conduct, analysis, and reporting. BMJ. 2011;342(apr11 1):d1766–d1766. doi:10.1136/bmj.d1766

23. Steichele K, Keefer A, Dietzel N, Graessel E, Prokosch HU, Kolominsky-Rabas PL. The effects of exercise programs on cognition, activities of daily living, and neuropsychiatric symptoms in community-dwelling people with dementia—a systematic review. Alzheimers Res Ther. 2022;14(1):1–13.

24. Harwood RH, Goldberg SE, Brand A, et al. Promoting Activity, Independence, and Stability in Early Dementia and mild cognitive impairment (PrAISED): randomised controlled trial. bmj. 2023;382. Accessed October 10, 2023. https://www.bmj.com/content/382/bmj-2023-074787

25. Position statement on use of the EQ-5D-5L value set for England (updated October 2019) | Technology appraisal guidance | NICE guidance | Our programmes | What we do | About. NICE. Accessed April 13, 2023. https://www.nice.org.uk/about/what-we-do/our-programmes/nice-guidance/technology-appraisal-guidance/eq-5d-5l

26. Health state life expectancies, UK – Office for National Statistics. Accessed April 13, 2023. https://www.ons.gov.uk/peoplepopulationandcommunity/healthandsocialcare/healthandlifeexpectancies/bulletins/healthstatelifeexpectanciesuk/2018to2020

27. Rubin DB. Multiple Imputation for Nonresponse in Surveys. Vol 81. John Wiley & Sons; 2004.

28. Simons CL, Rivero-Arias O, Yu LM, Simon J. Multiple imputation to deal with missing EQ-5D-3L data: Should we impute individual domains or the actual index? Qual Life Res. 2015;24(4):805–815.

29. Karen Jones, Amanda Burns. Unit Costs of Health and Social Care 2021 | PSSRU. Accessed February 23, 2023. https://www.pssru.ac.uk/project-pages/unit-costs/unit-costs-of-health-and-social-care-2021/

30. Curtis LH, Burns AT. Unit Costs of Health and Social Care 2020 | PSSRU. Accessed November 15, 2022. https://www.pssru.ac.uk/project-pages/unit-costs/unit-costs-2020/

31. Client Service Receipt Inventory (CSRI) | Client Service Receipt Inventory. Accessed April 14, 2023. https://www.pssru.ac.uk/csri/client-service-receipt-inventory/

32. Economic evaluation | NICE health technology evaluations: the manual | Guidance | NICE. Published January 31, 2022. Accessed April 13, 2023. https://www.nice.org.uk/process/pmg36/chapter/economic-evaluation

33. Briggs AH, Gray AM. Handling uncertainty in economic evaluations of healthcare interventions. Bmj. 1999;319(7210):635–638.

34. Johansson MM, Marcusson J, Wressle E. Cognitive impairment and its consequences in everyday life: experiences of people with mild cognitive impairment or mild dementia and their relatives. Int Psychogeriatr. 2015;27(6):949–958.

35. Organization WH. Mental Health and COVID-19: Early Evidence of the Pandemic’s Impact: Scientific Brief, 2 March 2022. World Health Organization; 2022.

36. Husereau D, Drummond M, Augustovski F, et al. Consolidated health economic evaluation reporting standards (CHEERS) 2022 explanation and elaboration: a report of the ISPOR CHEERS II Good Practices Task Force. Value Health. 2022;25(1):10–31.

37. Vemer P, Ramos IC, Van Voorn GAK, Al MJ, Feenstra TL. AdViSHE: a validation-assessment tool of health-economic models for decision makers and model users. Pharmacoeconomics. 2016;34(4):349–361.

38. Ho AD, Yu CC. Descriptive statistics for modern test score distributions: Skewness, kurtosis, discreteness, and ceiling effects. Educ Psychol Meas. 2015;75(3):365–388.

39. Doungsong K, Hartfiel N, Gladman J, Harwood RH, Edwards RT. RCT-based Social Return on Investment (SROI) of a home exercise programme for people with early dementia comparing in-person and blended delivery before and during the COVID-19 pandemic. medRxiv. Published online 2023:2023-08. Accessed October 23, 2023. https://www.medrxiv.org/content/10.1101/2023.08.25.23294408.abstract

